# Chagas Disease Screening: Awareness, Practices and Perceived Barriers Among Health Care Providers in Connecticut

**DOI:** 10.1101/2024.09.30.24314661

**Authors:** Erica J. Rayack, Maria Alejandra Gutierrez-Torres, Bernardo Lombo, Helen Mahoney-West, Sten Vermund

**Affiliations:** Yale School of Public Health, Yale University, New Haven, Connecticut, United States of America; Yale School of Medicine, Yale University, New Haven, Connecticut, United States of America; School of Medicine, Universidad de Los Andes, Bogota, Colombia; Boston Children’s Hospital, Boston, Massachusetts, United States of America

## Abstract

**Background:** An estimated 288,000 people in the United States are infected with Chagas disease. Despite published recommendations for healthcare providers, the neglected nature of this disease persists, in part due to gaps in knowledge and low levels of Chagas disease awareness among healthcare providers. A survey was emailed to healthcare providers to assess knowledge of Chagas disease, screening practices, and provider-perceived barriers to screening.

**Methods:** The survey link was emailed to healthcare providers and department heads. Providers answered a series of questions categorized either as knowledge (K), practice (P), or perceived barriers (B), which were then scored by category. The analysis included Spearman’s rho test to measure the strength of the correlation coefficient between K and P scores and between K and B scores, Kruskal-Wallis tests to see if average K, P, and B scores were equal across specialty groups, and Pairwise Wilcoxon Rank Sum Tests to assess for a significant difference between average scores of cardiology, infectious disease, and the “other specialties” category.

**Results:** 92 providers consented to complete the survey, 88 of whom fit the inclusion criteria of either affiliation with Yale University or YNHH. Infectious disease scored highest on average in knowledge and practices and reported lower provider-perceived barriers (0.66±0.183, 0.35±0.247, 0.42±0.365). No specialty received higher than 0.66±0.183 on average for knowledge or screening practices. Average perceived barriers to screening were highest in the “other” specialty category (0.76±0.303). There was a significant positive association between knowledge scores and screening practice scores and a significant negative association between knowledge scores and perceived barriers to screening.

**Conclusions:** This study revealed a knowledge deficit among healthcare providers in various specialties, which was associated with less frequent adherence to recommended screening practices for Chagas disease, as well as higher provider-perceived barriers to screening. This information can inform provider education initiatives so that patients at risk for Chagas disease may be better served in Connecticut and throughout the United States.

## Introduction

Chronic Chagas disease infections in the U.S. are most prevalent in populations who have emigrated from endemic areas of Latin America (Forsyth et al., 2022; Pérez-Molina & Molina, 2018). The ongoing need to spread awareness and improve screening must be considered within the context of xenophobia and stigma surrounding diseases associated with poverty (Forsyth et al., 2021; Montgomery et al., 2014). Some call for further analysis of “the political and economic structures that cause a widespread, life-threatening disease to persist in the shadows” (Forsyth et al., 2021). Others call for increased advocacy to raise awareness of Chagas disease (Ayres et al., 2022). Many specifically focus on the deficits in knowledge among healthcare providers and the outcomes related to these knowledge gaps (Amstutz-Szalay, 2017; Edwards et al., 2018; Mahoney- West et al., 2021, 2022; Stimpert & Montgomery, 2010; Verani et al., 2010).

Little is known about provider awareness and screening practices in Connecticut. The counties of Fairfield, New Haven, and Hartford have the state’s highest Hispanic populations (21.4%, 19.7%, 18.5%) (U.S. Census Bureau, 2021). As of 2021, the majority of Latin America-born immigrants living in Connecticut were from Mexico, Brazil, Colombia, and El Salvador (4.2%, 3.9%, 3.7%, 1.4%) (Migration Policy Institute, n.d.). Though these are all endemic countries, country prevalence may be an unreliable predictor of prevalence among U.S. immigrants due to within-country heterogeneity (Irish et al., 2022).

New Haven County is particularly interesting due to its connection to an extensive healthcare system and academic institutions. A survey was emailed to physicians and advanced practice providers (NPs, CNMs, PAs) affiliated with Yale-New Haven Hospital (YNHH) and/or Yale University. Data were collected on healthcare provider knowledge and practices surrounding Chagas disease, as well as provider-perceived barriers to screening. It was predicted that there would be an overall lack of awareness surrounding Chagas disease and its risks. This lack of knowledge would be associated with less frequent screening practices and higher provider-perceived barriers to screening. This was based on results seen in similar provider survey studies (Amstutz-Szalay, 2017; Mahoney-West et al., 2021; Stimpert & Montgomery, 2010). Additionally, among providers, cardiology and infectious disease specialists were expected to be significantly more knowledgeable about Chagas disease, score significantly higher on the practice category questions, and report significantly lower perceived barriers to Chagas disease screening. This survey was an important step toward measuring adherence to professional recommendations and fostering equitable patient care.

## Methods

This study was granted an exemption by the Yale Institutional Review Board (IRB# 2000033998). Consent to participate was asked as the first question on the RedCap survey. A stop action was applied to the survey, halting participation for anyone who did not consent.

### Survey and Scoring

Recruitment took place from January 2, 2023, to April 7, 2023. Research Electronic Data Capture (REDCap) software was used to create the survey instrument and link and capture respondent data. The survey link was shared via email with healthcare providers who are affiliated with Yale University or YNHH. The email containing the survey explanation and link is in Appendix A. The survey instrument is in Appendix B. Participants were recruited through public YNHH directories, networks at the author’s affiliated academic institutions (YSN and YSPH), and through emails by department leaders and administrators.

Providers answered a series of questions categorized either as knowledge (K), practice (P), or perceived barrier (B). There were nine knowledge questions, three practice questions, and three perceived barrier questions. Targeted provider specialties included the following nine specialty categories relevant to Chagas disease screening: Primary Care/Family/Internal Medicine, Pediatric Primary Care, Women’s Health/Midwifery/OBGYN, Cardiology (Adult), Cardiology (Pediatric), Gastroenterology (Adult), Gastroenterology (Pediatric), Infectious Disease (Adult), Infectious Disease (Pediatric). All other respondents were sorted into the category “other.” Due to low response rates within certain specialties, the “other” category was later expanded to all aside from the main categories of interest, cardiology, and infectious disease (**Table 1**).

**Table 1.**
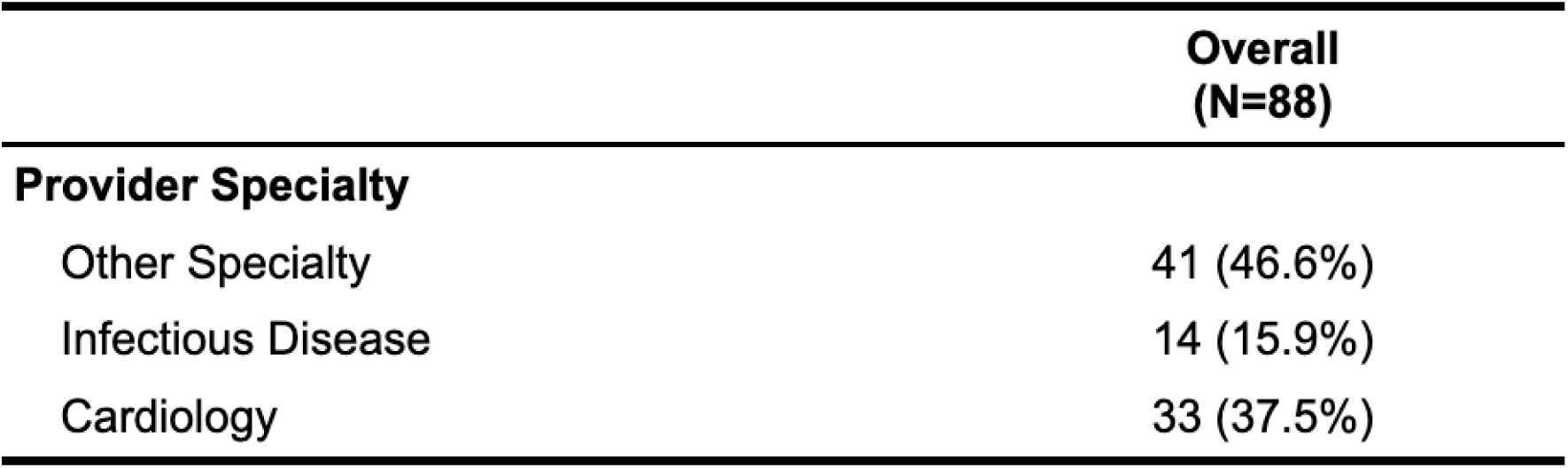
Participating providers categorized by Other, Infectious Disease, and Cardiology.

Respondents received a score for each category (K, P, and B). All knowledge questions were scored out of 1 point, with correct answers to yes or no questions and self-reported confidence in knowledge/familiarity resulting in a full point for that K question. One K question was originally 2 points total but was weighted accordingly so as not disproportionately to affect the participant’s K score. A “check all that apply” question (#10) was categorized as correct (1 point) only if all four correct options were selected. Near-correct answers (2 out of 4 or 3 out of 4) did not contribute to the participant’s total knowledge score. For each question regarding provider practices (P), participants were presented with a choice between “Never,” “Rarely,” “Sometimes,” “Frequently,” and “Always.” These questions were scored ordinally as follows: Never (0), Rarely (1), Sometimes (2), Frequently (3), Always (4). Each P question was out of 4 points; no weighted calculation was required. Higher scores P indicated more frequent Chagas disease screening. Perceived barrier questions were scored ordinally as follows: Not a barrier at all (0), Minor barrier (1), Moderate barrier (2), Major barrier (3). Each B question was out of 3 points, and no weighted calculation was required. Higher B scores indicated higher barriers to screening perceived by the individual clinician.

Statistical Analysis

R statistical software (version 4.2.3) was used for data analysis. K, P, and B scores were based on the proportion of correct answers in each category. Scores were grouped by provider specialty, and mean scores by specialties were calculated (**Figure 1a-c**). Spearman’s rho test was used to measure the correlation strength between mean K and P scores and between mean K and B scores (**Figure 2a-b**). Kruskal-Wallis tests were used to check for equivalence in K, P, and B scores across specialty groups. This non-parametric test was selected after Shapiro–Wilk Tests for K, P, and B scores yielded p-values below 0.05 (see histograms in supplemental figures for visualization of skew). This was followed by Pairwise Wilcoxon Rank Sum Tests with the Bonferroni p-value adjustment method to assess for a difference between cardiology and infectious disease.

**Figure 1.**
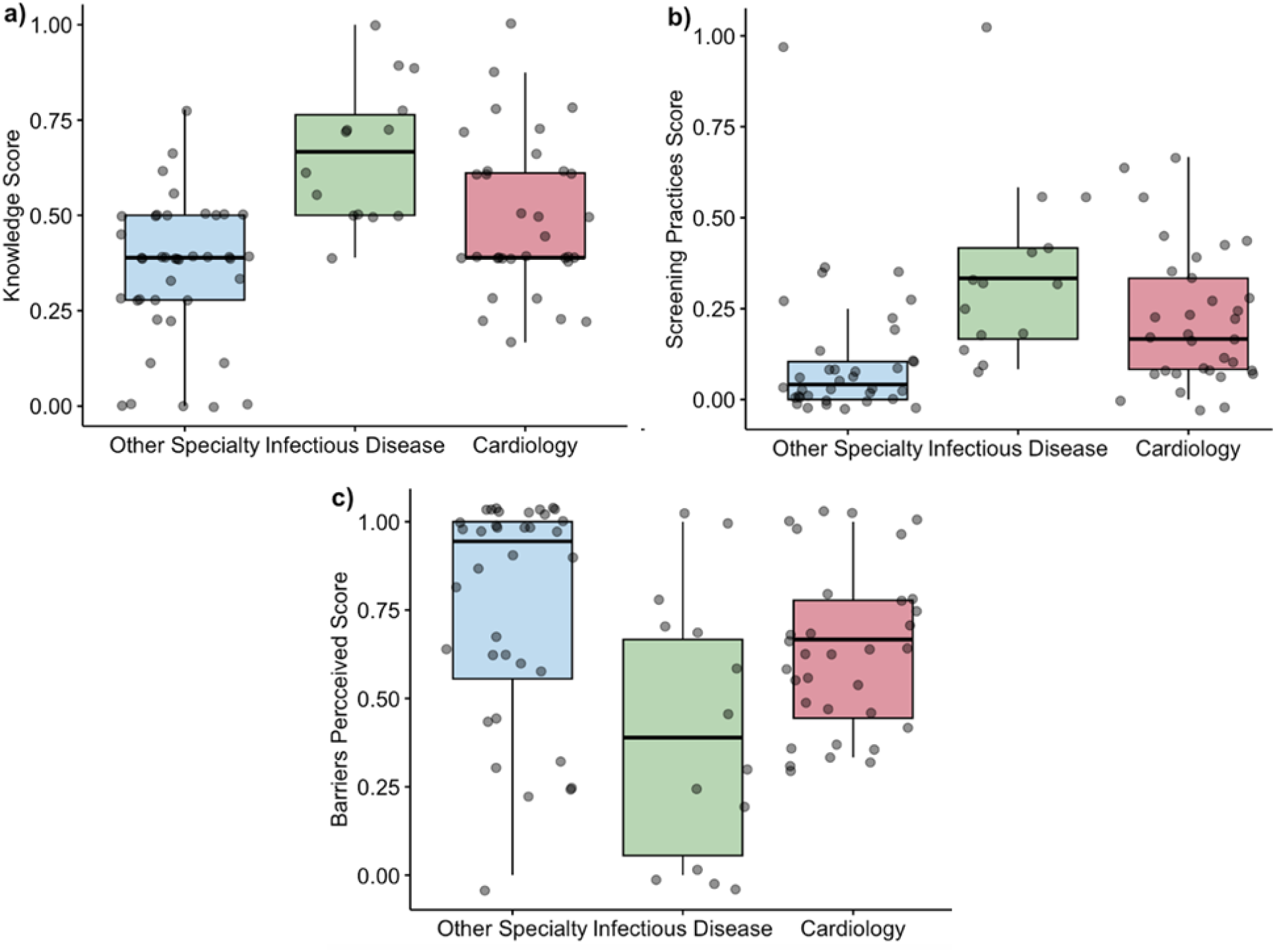
Boxplots by specialty show a) knowledge, b) screening practices, and c) perceived barriers.

**Figure 2.**
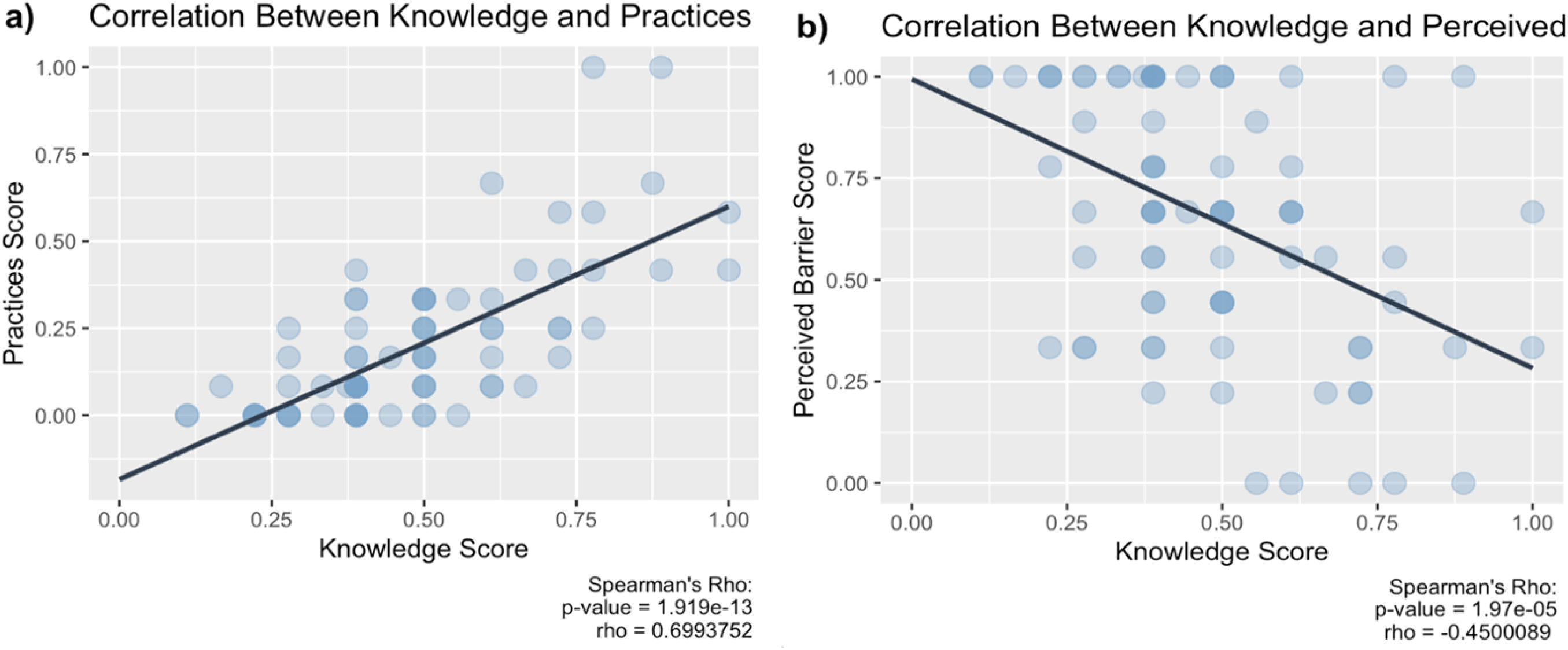
a) Correlations between K and P scores. K and B scores, with results of Spearman’s rank correlation coefficient shown.

## Results

92 providers consented to complete the survey, 88 of whom fit the inclusion criteria of affiliation with Yale University and/or YNHH. Counts and percentages of provider affiliation, provider type, and provider specialty can be found in **Table 2**.

**Table 2.**
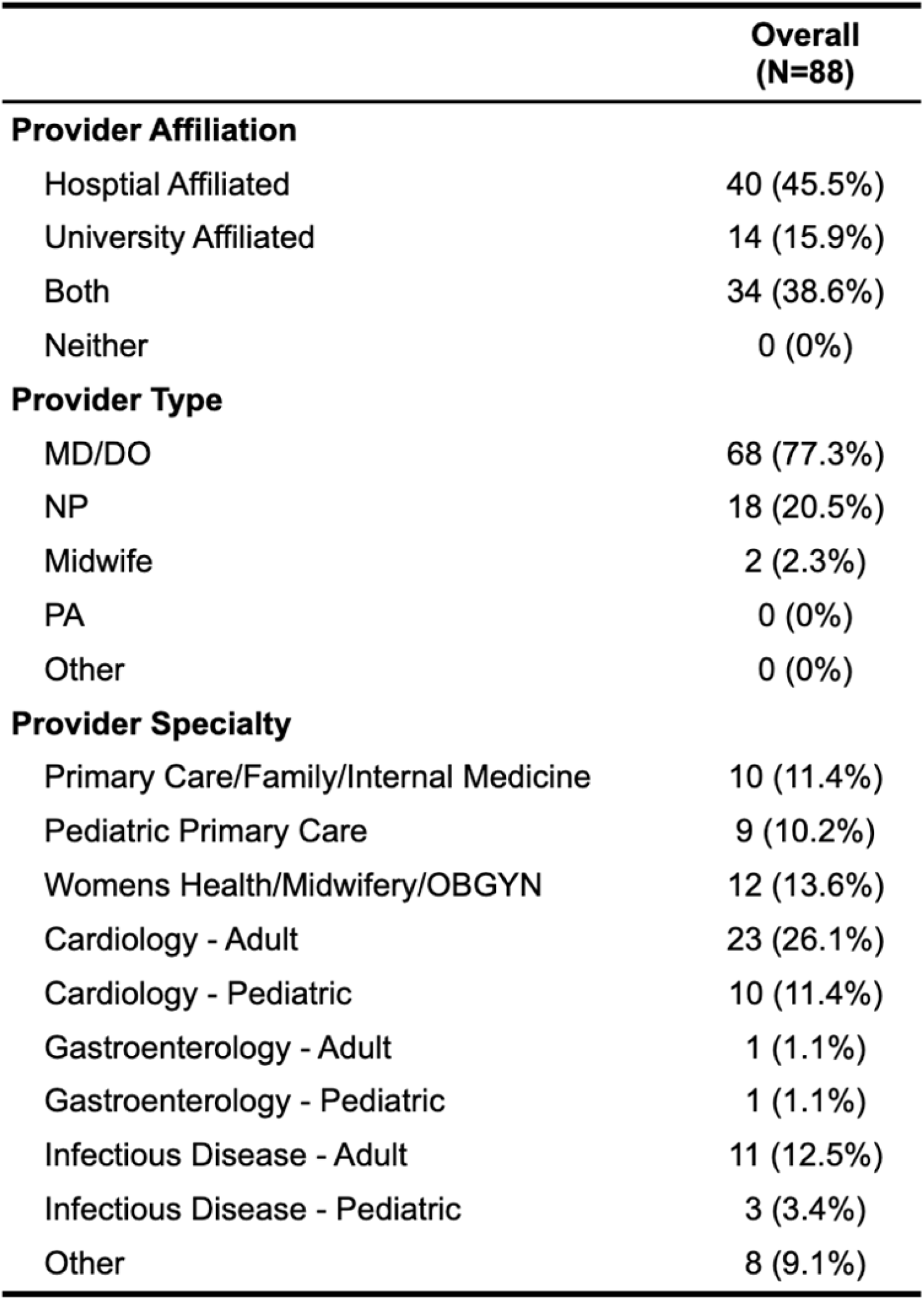
Participating providers are categorized by affiliation, type, and specialty.

Five of the 88 respondents had never heard of Chagas disease, resulting in the automatic termination of the survey. All five providers unaware of Chagas disease were nurse practitioners (1 Women’s Health, 2 Pediatric primary care NPs, 1 Primary Care NP, and 1 “other” specialty), resulting in 75% of NPs/Midwives having heard of Chagas disease. These individuals received a 0 for their total knowledge scores and “not applicable” for all practice and barrier questions. Boxplots of provider scores by specialty can be found in **Figure 1a-c**. 100% of MD/DO respondents had heard of Chagas disease. Infectious disease scored highest on average in K and P and reported lower provider-perceived barriers to screening patients (0.66±0.183, 0.35±0.247, 0.42±0.365). No specialty received higher than 0.66±0.183 on average for knowledge or screening practices. Average perceived barriers to screening were highest in the “other” specialty category (0.76±0.303). Spearman’s rho test revealed a positive association between K and P (p <.0001, rho = 0.70) and a negative association between K and B (p-value <.0001, rho = -0.45) (**Figure 2a-b**). Due to the fact that the normality assumption was not held (Se supplemental section), K, P, and B scores were compared across the three specialty groups using the Kruskal-Wallis rank sum test. Knowledge scores, screening practices scores, and the perceived barrier scores differed by specialty category. This was followed by three Pairwise Wilcoxon Rank Sum Tests with Bonferroni p-value adjustments to investigate these differences across specialty categories further. Though an underpowered survey, as is detailed further in the discussion, the interpretation of the results was as follows.

Regarding knowledge scores, infectious disease specialist scores were greater than both cardiology (p=.02) and “other” specialties (p<.0001). Cardiology knowledge scores were greater than the “other” specialties scores (p=0.052). Screening practice scores for both infectious disease and cardiology were higher than “other” specialties (p<0.001, p<0.005), but infectious disease and cardiology practice scores did not show a stark difference (p=0.17). Perceived barrier scores of infectious disease and cardiology were lower than “other” (p=0.01, p=0.08), and infectious disease scores did not reveal a difference from cardiology (p=0.16).

## Discussion

Though Chagas disease is likely underdiagnosed within the United States, efforts to estimate prevalence, diagnose at-risk populations, and treat have gradually increased (Forsyth et al., 2021; Ikedionwu et al., 2020). Despite publications providing screening and diagnosis recommendations, this disease remains neglected (Forsyth et al., 2022; PAHO, 2019). An estimated 288,000 people in the U.S. are infected, with chronic infections making up the vast majority and acute infections seen only sporadically (Carter et al., 2012; Irish et al., 2022). Of these 288,000, an estimated 57,000 have Chagas cardiomyopathy (Irish et al., 2022). To obtain prevalence estimates among Latin America-born U.S. residents, Irish et al., 2022 relied upon U.S. *T. cruzi* seroprevalence data to the extent it was available. Beyond this, the authors used 2014–2018 microdata from IPUMS-USA—a database built from federal censuses and American Community Surveys—in addition to World Health Organization reports on seroprevalence in disease-endemic countries (Irish et al., 2022). These updated estimates are similar to those previously calculated but are likely more accurate due to relative improvements in U.S. seroprevalence data (Bern & Montgomery, 2009; Irish et al., 2022; Manne-Goehler et al., 2016).

Chronic infection can be indeterminate or determinate, with indeterminate signifying antibody seropositivity but normal electrocardiogram findings, standard chest x-rays, and the absence of digestive mega syndromes (Rassi et al., 2010). Of those with chronic Chagas disease, approximately 30-40% develop determinate (symptomatic) Chagas disease, which can manifest in various ways, including cardiac conduction abnormalities, cardiomyopathy, megaesophagus, and megacolon (Pérez-Molina & Molina, 2018; Rassi et al., 2010). Cardiac involvement can eventually result in heart failure and sudden death (Pérez-Molina & Molina, 2018).

An estimated 43,000 women of reproductive age in the U.S. are infected with Chagas disease, and, as a result, approximately 22-108 congenitally infected infants are born in the U.S. each year (Irish et al., 2022). Experts on this life-threatening disease urgently call for updated guidelines for cardiologists and obstetrics/gynecology specialists (Ayres et al., 2022). The public health importance of screening in the U.S. is highlighted by the reduced risk of vertical transmission in those treated prior to pregnancy and the >90% cure rate in infants treated within the first year of life (Carlier et al., 2019; Forsyth et al., 2022). Not only has treatment prior to pregnancy been found even in mothers who had a previous occurrence of vertical transmission. (Murcia et al., 2017; Sosa-Estani et al., 2009). Once treated and PCR negative, no congenital Chagas disease occurred with subsequent pregnancies. In the larger at-risk population, more attention to screening, diagnosis, and treatment may lead to better patient outcomes, more appropriate interventions, and better U.S. prevalence estimates (Ayres et al., 2022; Crespillo-Andújar et al., 2022; Edwards et al., 2017; Forsyth et al., 2022; Meymandi et al., 2018; PAHO, 2019).

The causative agent of Chagas disease is the hemoflagellate, *Trypanosoma cruzi*, a protozoan parasite that is spread to humans primarily by the triatomine insect. Triatomines, colloquially called “kissing bugs,” are in the order Hemiptera, family Reduviidae, and subfamily Triatominae. This nocturnal insect colonizes around and inside homes, both in Latin America and in the Southwest U.S. (Alvarado et al., 2022; Dye-Braumuller et al., 2020; Forsyth et al., 2022; Klotz et al., 2021; Klotz & Schmidt, 2020; Rassi et al., 2010). Historically, transmission from insect vector to human host was attributed predominantly to the vector taking a blood meal and defecating near the bite punctum, after which host behaviors, like scratching or rubbing, are thought to facilitate the spread of parasite-infested feces into the host bloodstream (Klotz & Schmidt, 2020; Rassi et al., 2010). Infections also spread through oral ingestion, vertical transmission, and blood/organ donation (Rassi et al., 2010). Additionally, transplant-related immunosuppression has been associated with Chagas disease reactivation in chronically infected recipients (Hamilton et al., 2022; Radisic & Repetto, 2020).

The oral route of transmission has received increased attention due in part to notable foodborne outbreaks observed in South America and reports of triatomines in kitchens and food containers within the U.S. (Alarcón de Noya et al., 2015; FAO/WHO, 2014; Klotz & Schmidt, 2020; Nóbrega Food and Agriculture Organization of the United Nations (FAO) has ranked *T. cruzi* 10^th^ out of the 24 most important foodborne parasites globally. However, foodborne transmission is thought to be primarily relevant to endemic countries (FAO/WHO, 2014). *T. cruzi* does not replicate in food, so oral transmission depends instead on contamination, such as vector fecal matter in fruit juices or ingestion of parasite-infected reservoir animals (FAO/WHO, 2014). The dominant mode of autochthonous transmission within the U.S. remains undetermined, though oral transmission has been proposed (Klotz & Schmidt, 2020). This suggestion is supported by triatomine defecation indices and a lack of association between U.S. bite reports and positive serological findings (Klotz & Schmidt, 2020).

Despite its critical relevance in diagnosing and managing diseases like Chagas, the diminishing focus on parasitology in medical education poses a significant barrier to reducing disease burden. Inadequate representation of parasitology in medical curricula contributes to the scarcity of healthcare professionals equipped to tackle such diseases. This troubling trend is reflected in well-established medical texts like Harrison’s Principles of Internal Medicine, which reduced coverage of parasitic diseases from sixty pages in the 1950s to four pages by 2002.(Meador, 2004; Melendez, 2003). This reduction is mirrored globally, with a decreased presence of medical faculty specializing in parasitology across Europe, Japan, and China (Bruschi, 2009; Sekine, 2022; Peng et al., 2012). The rapid retirement of parasitology experts exacerbates this gap in expertise (Barnish et al., 2006).

Despite recent publications providing clear recommendations for screening and diagnosis, it is not surprising that the neglected nature of this disease persists (CDC, 2023; Forsyth et al., 2022; PAHO, 2019). Not only is it a disease historically associated with rural poverty, but, at least within 2022; Mahoney-West et al., 2021). Additionally, it is a silent, sometimes decades-long disease process (Forsyth et al., 2021; Hotez et al., 2013; Pérez-Molina & Molina, 2018). With disease characteristics such as this, the efforts toward increasing Chagas disease education for healthcare providers are critical to further understanding and lessening the burden of this disease within the U.S.

In this study, knowledge of Chagas disease was strongly linked to better screening practices and lower provider-perceived barriers to screening at-risk patients. The general lack of knowledge, infrequent screening practices, and perception of high barriers to screening are not unique to this study population (Amstutz-Szalay, 2017; Edwards et al., 2018; Mahoney-West et al., 2021; Malhotra et al., 2021; Soares Cajaiba-Soares et al., 2021; Stimpert & Montgomery, 2010; Verani et al., 2010). The small respondent size was a notable study limitation. This should be considered when interpreting provider scores, as reaching the significance threshold of p<0.05 may be due to chance rather than the actual difference between specialties or the true relationship between variables.

Education sessions would likely improve provider adherence to recommendations and provide them with the tools to serve at-risk Latin American patients better (Mahoney-West et al., 2022). Though infectious disease specialists and cardiologists had higher screening practice scores, these specialists likely see patients after symptomatic disease has developed, be it acute or chronic symptomatic, depending on the specialty. In the survey instrument, practice questions focused on screening, but questions did not clearly specify whether this meant asymptomatic screening, as was pointed out by one participant. Asymptomatic screening practices compared to diagnostic testing practices may be a beneficial area to explore in future research. Improvement in screening OB-GYN providers is warranted, especially when the importance of preventing cases of vertical transmission is considered (Alarcón et al., 2016; CDC, 2012; Forsyth et al., 2022).

## Study limitations

This study examines the level of knowledge and screening practices related to Chagas disease among healthcare providers in Connecticut. Although the study has certain limitations, it offers valuable insights and perspectives. The response rate, although modest, encompassed a diverse range of specialties, indicating a widespread interest in Chagas disease within various medical domains. On the other hand, although self-reported data may be subject to bias, such as social desirability effects, it provides valuable information on providers’ perceived knowledge and practices. This information is essential for gaining a comprehensive understanding of the current challenges in screening behaviors. Thirdly, the findings primarily pertain to a single institution and may not be generalizable to other contexts. However, they do provide valuable and specific insights that could be relevant in similar healthcare settings. The study’s cross-sectional design limits our ability to establish causal relationships between knowledge and practices. However, it effectively highlights current associations that can provide valuable insights for targeted educational efforts. Also, the intricacy of the scoring system, while it may result in misinterpretations, offers comprehensive data that can precisely identify areas in need of enhancement. Subsequent studies could enhance these discoveries by utilizing a larger and more varied group of participants, as well as adopting a longitudinal methodology to more accurately observe patterns and establish cause and effect relationships in relation to provider practices regarding Chagas disease.

## Acknowledgments

Our sincere gratitude to Dr. Sangchoon Jeon for your generosity with your time and guidance on statistical analysis and to Dr. Amy Bei for your wisdom and encouragement on this project.

## Data availability

All the data is publicly available without restrictions upon reasonable request to the authors.

## Supplementary Materials

**5. Supplementary Material 1:** Email to Lead Department Contact

**2. Supplementary** Figure 1: Histograms showing the distribution of all participants’ knowledge scores, practice scores, and perceived barriers scores.

**3. Supplementary Table 1:** Data Dictionary Codebook

**6. Supplementary Material 1:** Email to Lead Department Contact

Email Subject:

Health Care Provider Survey To Complete

Email Body:

Dear Lead Department Contact

Thank you for your willingness to distribute this survey within your clinic/medical department. This study was granted an IRB exception (IRB Protocol ID is 2000033998). Please email the below message to all providers on your clinic/medical department contact list:

<u>Awareness, Practices, and Perceived Barriers of a Neglected Disease – Provider Survey</u>

Please complete this 5-minute <u>survey</u>. Every response is valuable, no matter the knowledge level. The survey is entirely anonymous and investigates provider awareness, practices, and perceived barriers to Chagas disease screening. It will be used to assess the need for education and communication regarding this neglected disease.

This study was granted IRB exemption (IRB Protocol ID: 2000033998). If you have any questions, please.

If the above survey link does not work, please copy and paste the following into your address bar:

Thank you for your time,

**Erica J. Rayack** (she/they)

Joint Degree MSN & MPH Student Yale School of Nursing

Yale School of Public Health

**Supplementary Table 1:**
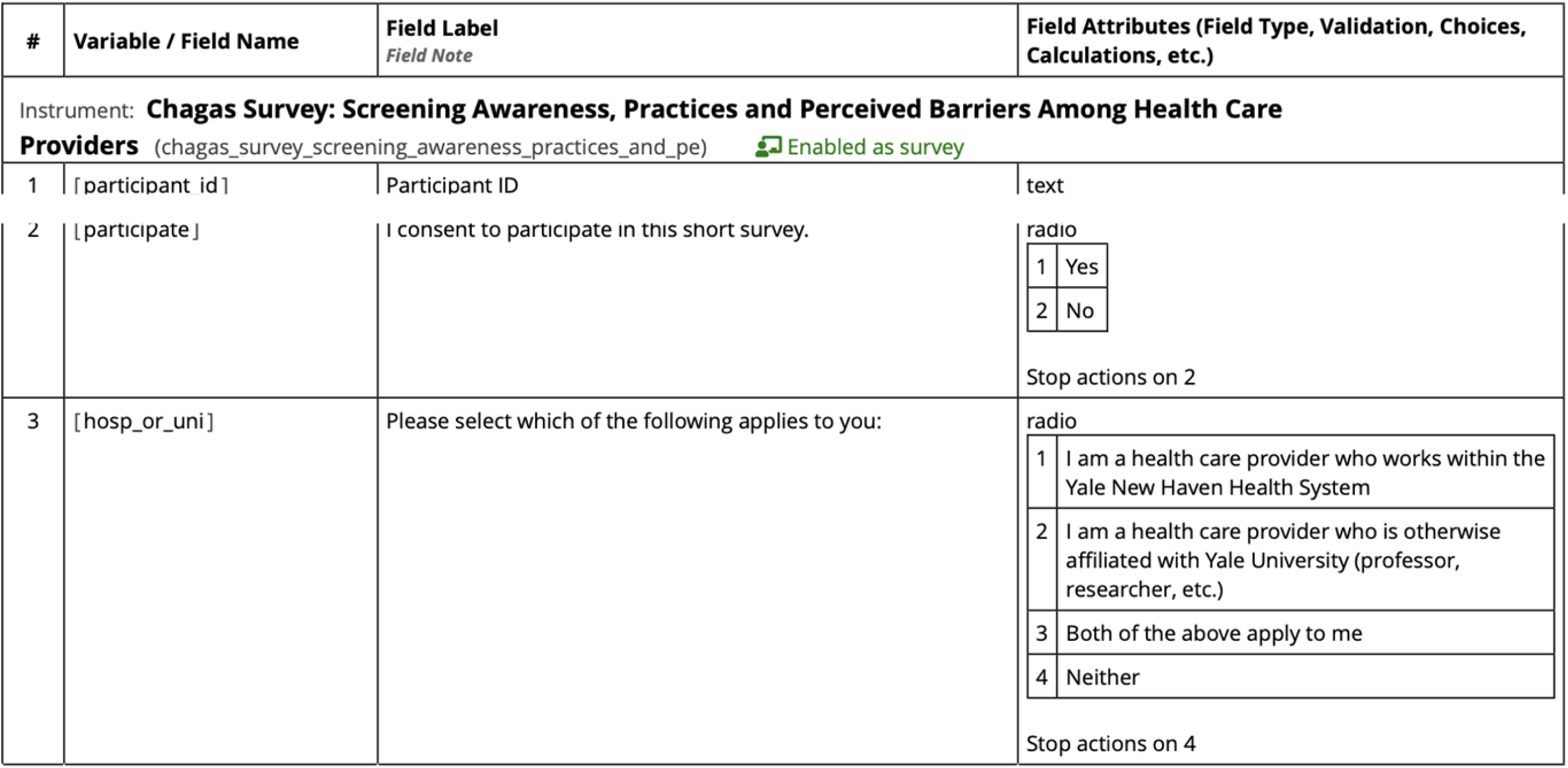
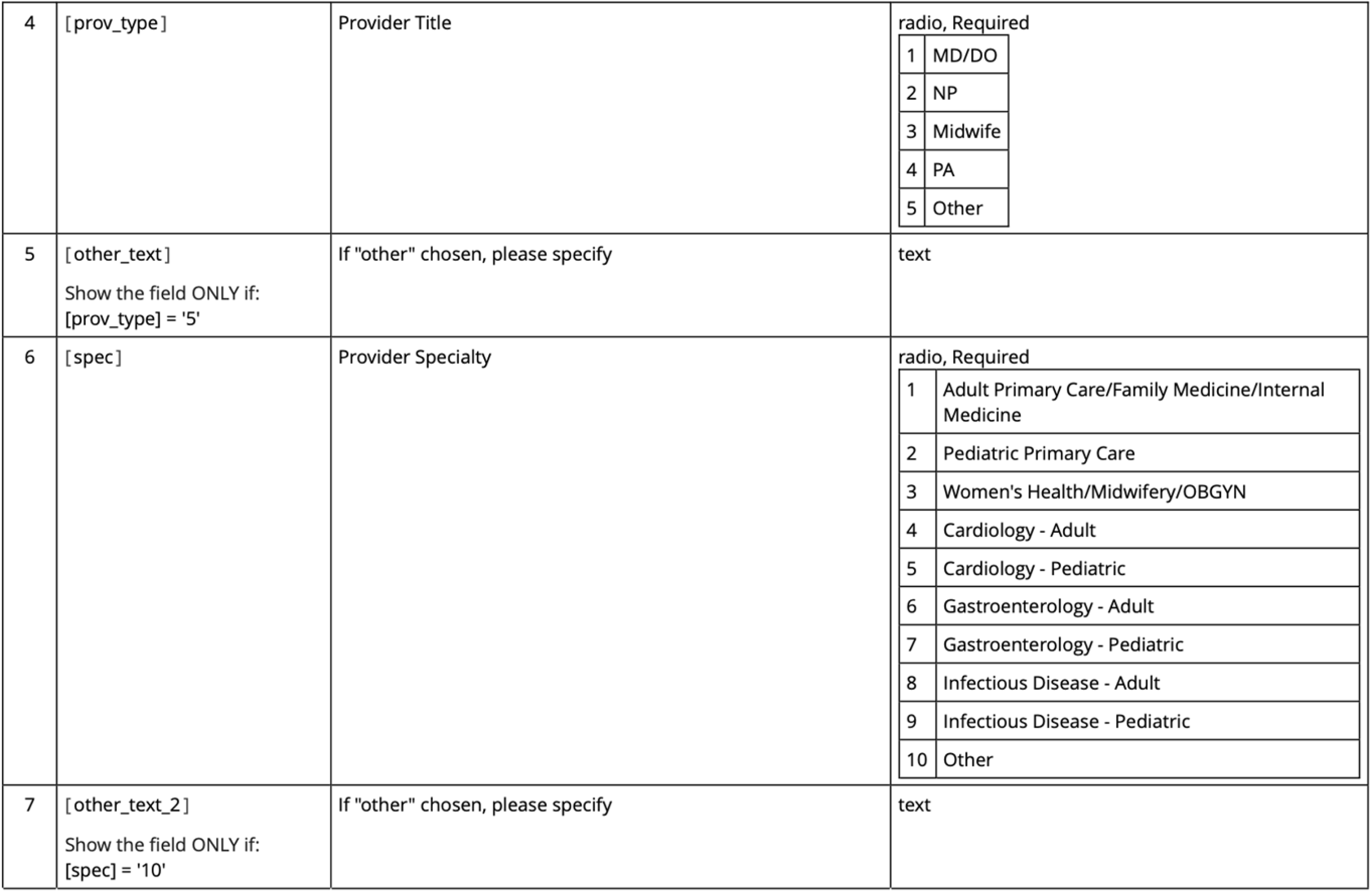
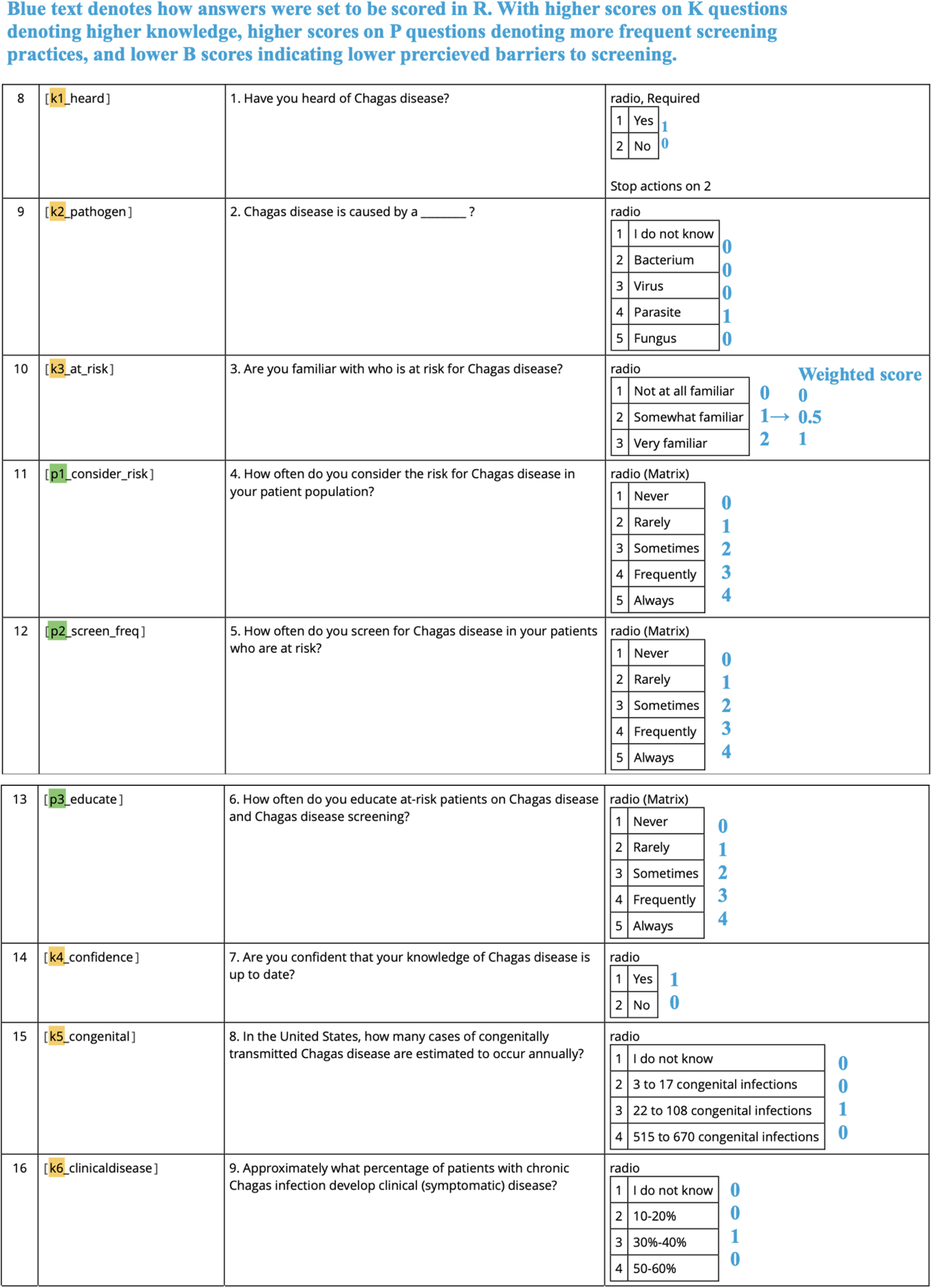
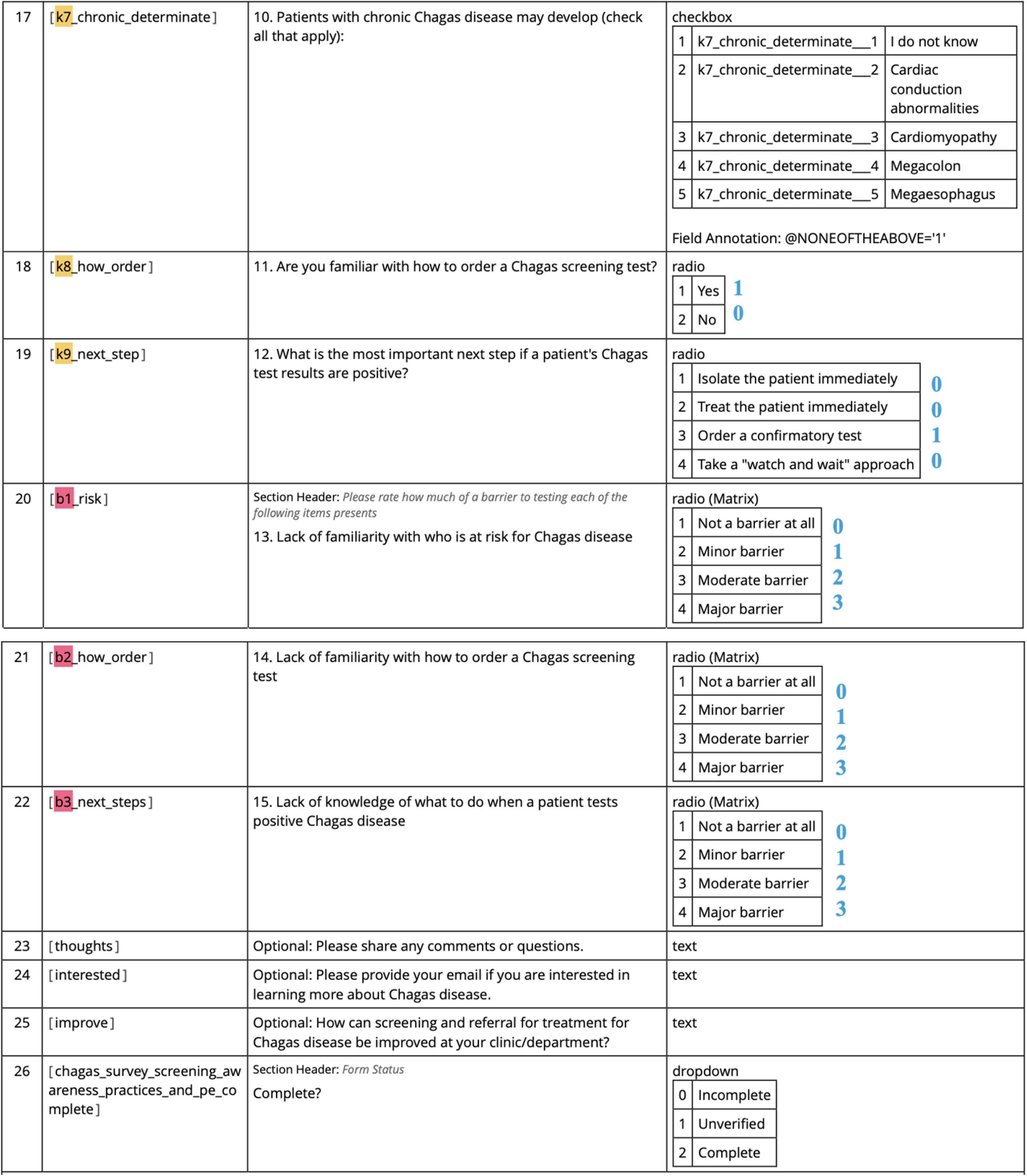
Data Dictionary Codebook.

**Supplementary Figure 1:**
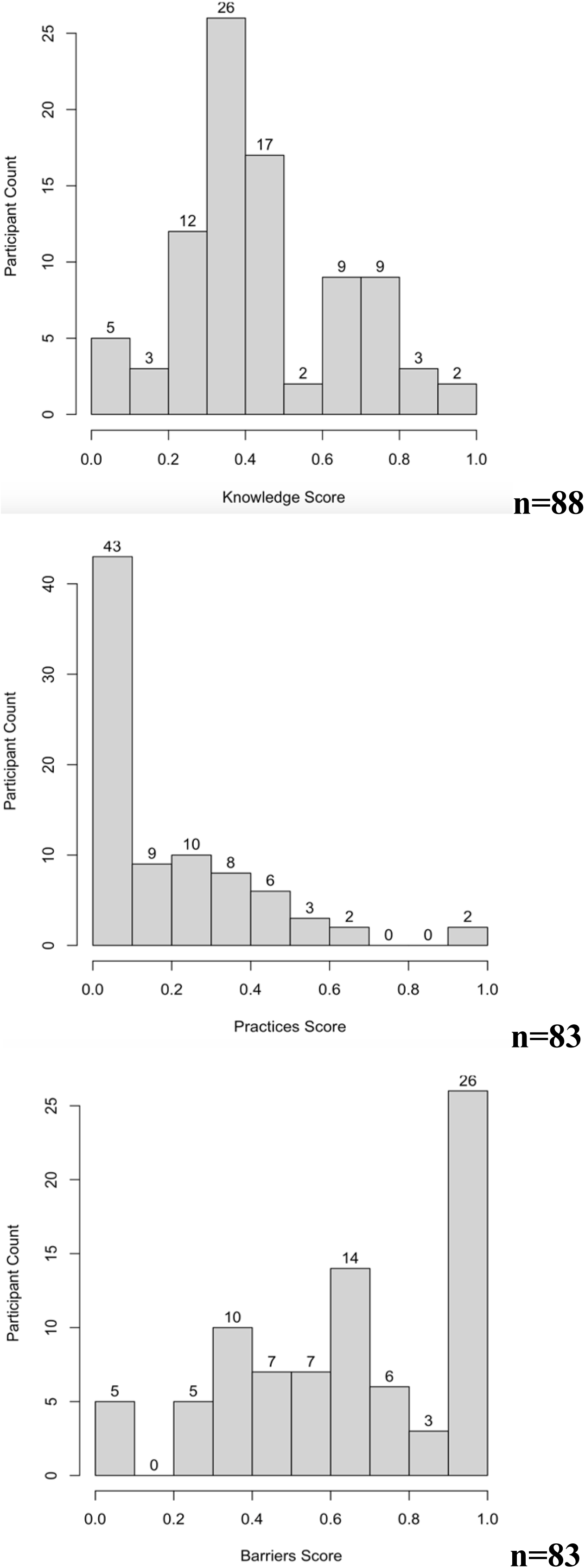
Histograms showing the distribution of all participants’ knowledge scores, practice scores, and perceived barriers scores.

## REFERENCES

1. Acholonu ADW (2003). Trends in teaching parasitology: the American situation. Trends Parasitol 19:6–9

2. Alarcón, A., Morgan, M., Montgomery, S. P., Scavo, L., Wong, E. C. C., Hahn, A., & Jantausch, B. (2016). Diagnosis and Treatment of Congenital Chagas Disease in a Premature Infant. Journal of the Pediatric Infectious Diseases Society, 5(4), e28–e31. 10.1093/jpids/piw043

3. Amstutz-Szalay, S. (2017). Physician Knowledge of Chagas Disease in Hispanic Immigrants Living in Appalachian Ohio. Journal of Racial and Ethnic Health Disparities, 4(3), 523–528. 10.1007/s40615-016-0254-8

4. *Angheben*, *A.*, *Boix*, *L.*, *Buonfrate*, *D.*, *Gobbi*, *F.*, *Bisoffi*, *Z.*, *Pupella*, *S.*, *Gandini*, *G.**, &* *Aprili*, *G*. (2015). Chagas disease and transfusion medicine: a perspective from non-endemic countries. Blood transfusion = Trasfusione del sangue, 13(4), 540–550. 10.2450/2015.0040-15

5. Ayres, J., Marcus, R., & Standley, C. J. (2022). The Importance of Screening for Chagas Disease Against the Backdrop of Changing Epidemiology in the USA. Current Tropical Medicine Reports, 9(4), 185–193. 10.1007/s40475-022-00264-7

6. Barnish, G., Crewe, W., & Theakston, R. D. (2006). Parasitologists lost? Trends in Parasitology, 22(10), 454–455. 10.1016/j.pt.2006.07.007

7. Bruschi F. (2009). How parasitology is taught in medical faculties in Europe? Parasitology, lost?. Parasitology research, 105(6), 1759–1762. 10.1007/s00436-009-1594-7

8. Carlier, Y., Altcheh, J., Angheben, A., Freilij, H., Luquetti, A. O., Schijman, A. G., Segovia, M., Wagner, N., & Albajar Vinas, P. (2019). Congenital Chagas disease: Updated recommendations for prevention, diagnosis, treatment, and follow-up of newborns and siblings, girls, women of childbearing age, and pregnant women. PLoS Neglected Tropical Diseases, 13(10), e0007694. 10.1371/journal.pntd.0007694

9. CDC. (2012). Congenital transmission of Chagas disease—Virginia, 2010. *MMWR*. Morbidity and Mortality Weekly Report. Centers for Disease Control and Prevention, 61(26), 477–479.

10. CDC. (2023, April 3). Chagas Disease—Resources for Health Professionals. https://www.cdc.gov/parasites/chagas/health_professionals/index.html

11. Clark, E. H., Marquez, C., Whitman, J. D., & Bern, C. (2022). Screening for Chagas Disease Should Be Included in Entry-to-Care Testing for At-Risk People With Human Immunodeficiency Virus (HIV) Living in the United States. Clinical Infectious Diseases, 75(5), 901–906. 10.1093/cid/ciac154

12. Despommier DD, Gwadz RW, Hotez PJ, Knirsch CA (2000) Preface. Parasitic Diseases, 4th ed. Apple Trees Productions, LLC, New York, pp viii–ix

13. Edwards, M. S., Abanyie, F. A., & Montgomery, S. P. (2018). Survey of Pediatric Infectious Diseases Society Members About Congenital Chagas Disease. The Pediatric Infectious Disease Journal, 37(1), e24–e27. 10.1097/INF.0000000000001733

14. Forsyth, C. J., Hernandez, S., Flores, C. A., Roman, M. F., Nieto, J. M., Marquez, G., Sequeira, J., Sequeira, H., & Meymandi, S. K. (2021). “You Don’t Have a Normal Life”: Coping with Chagas Disease in Los Angeles, California. Medical Anthropology, 40(6), 525–540. 10.1080/01459740.2021.1894559

15. Forsyth, C. J., Manne-Goehler, J., Bern, C., Whitman, J., Hochberg, N. S., Edwards, M., Marcus, R., Beatty, N. L., Castro-Sesquen, Y. E., Coyle, C., Stigler Granados, P., Hamer, D., Maguire, J. H., Gilman, R. H., Meymandi, S., & US Chagas Diagnostic Working Group. (2022). Recommendations for Screening and Diagnosis of Chagas Disease in the United States. The Journal of Infectious Diseases, 225(9), 1601–1610. 10.1093/infdis/jiab513

16. Hotez, P. J., Dumonteil, E., Betancourt Cravioto, M., Bottazzi, M. E., Tapia-Conyer, R., Meymandi, S., Karunakara, U., Ribeiro, I., Cohen, R. M., & Pecoul, B. (2013). An unfolding tragedy of Chagas disease in North America. PLoS Neglected Tropical Diseases, 7(10), e2300. 10.1371/journal.pntd.0002300

17. Irish, A., Whitman, J. D., Clark, E. H., Marcus, R., & Bern, C. (2022). Updated Estimates and Mapping for Prevalence of Chagas Disease among Adults, United States. Emerging Infectious Diseases, 28(7), 1313–1320. 10.3201/eid2807.212221

18. Lynn, M. K., Dye-Braumuller, K. C., Beatty, N. L., Dorn, P. L., Klotz, S. A., Stramer, S. L., Townsend, R. L., Kamel, H., Vannoy, J. M., Sadler, P., Montgomery, S. P., Rivera, H. N., & Nolan, M. S. (2022). Evidence of likely autochthonous Chagas disease in the southwestern United States: A case series of Trypanosoma cruzi seropositive blood donors. Transfusion, 62(9), 1808–1817. 10.1111/trf.17026

19. Mahoney-West, H., Milliren, C. E., Manne-Goehler, J., Davis, J., Gallegos, J., Perez, J. H., & Köhler, J. R. (2022). Effect of clinician information sessions on diagnostic testing for Chagas disease. PLOS Neglected Tropical Diseases, 16(6), e0010524. 10.1371/journal.pntd.0010524

20. Mahoney-West, H., Milliren, C. E., Vragovic, O., Köhler, J. R., & Yarrington, C. (2021). Perceived barriers to Chagas disease screening among a diverse group of prenatal care providers. PLOS ONE, 16(2), e0246783. 10.1371/journal.pone.0246783

21. Malhotra, S., Masood, I., Giglio, N., Pruetz, J. D., & Pannaraj, P. S. (2021). Current knowledge of Chagas-related heart disease among pediatric cardiologists in the United States. BMC Cardiovascular Disorders, 21(1), 116. 10.1186/s12872-021-01924-8

22. Meador C. K. (2004). From Med School: Shoes, Window Screens, and Meat. Medscape General Medicine, 6(2), 59.

23. Melendez RD (2003). Trends in teaching parasitology: Where to complain? Trends Parasitol 3:387

24. Migration Policy Institute. (n.d.). State Immigration Data Profiles—Connecticut. Migrationpolicy.Org. Retrieved April 19, 2023, from https://www.migrationpolicy.org/data/state-profiles/state/demographics/CT

25. Montgomery, S. P., Starr, M. C., Cantey, P. T., Edwards, M. S., & Meymandi, S. K. (2014). Neglected Parasitic Infections in the United States: Chagas Disease. The American Journal of Tropical Medicine and Hygiene, 90(5), 814–818. 10.4269/ajtmh.13-0726

26. Murcia, L., Simón, M., Carrilero, B., Roig, M., & Segovia, M. (2017). Treatment of Infected Women of Childbearing Age Prevents Congenital Trypanosoma cruzi Infection by Eliminating the Parasitemia Detected by PCR. The Journal of Infectious Diseases, 215(9), 1452–1458. 10.1093/infdis/jix087

27. Nunes, M. C. P., Beaton, A., Acquatella, H., Bern, C., Bolger, A. F., Echeverría, L. E., Dutra, W. O., Gascon, J., Morillo, C. A., Oliveira-Filho, J., Ribeiro, A. L. P., Marin-Neto, J. A., & null, null. (2018). Chagas Cardiomyopathy: An Update of Current Clinical Knowledge and Management: A Scientific Statement From the American Heart Association. Circulation, 138(12), e169–e209. 10.1161/CIR.0000000000000599

28. PAHO. (2019). Guidelines for the Diagnosis and Treatment of Chagas Diseases.

29. Peng, H. J., Zhang, C., Wang, C. M., & Chen, X. G. (2012). Current status and challenges of Human Parasitology teaching in China. Pathogens and global health, 106(7), 386–390. 10.1179/2047773212Y.0000000040

30. Pérez-Molina, J. A., & Molina, I. (2018). Chagas disease. The Lancet, 391(10115), 82–94. 10.1016/S0140-6736(17)31612-4

31. Schmunis GA, Yadon ZE. Chagas disease: a Latin American health problem becoming a world health problem. Acta Trop 2010; 115: 14-21.

32. Sekine, S. Pre-graduate teaching of human parasitology for medical laboratory technologist programs in Japan. Humanit Soc Sci Commun 9, 225 (2022). 10.1057/s41599-022-01246-w

33. Singh, A., Cohen, B., Sturzoiu, T., Vallabhaneni, S., & Shirani, J. (2020). Recent trends in hospital admissions and outcomes of cardiac Chagas disease in the United States. International journal of critical illness and injury science, 10(*3*), 134*–*139. 10.4103/IJCIIS.IJCIIS_85_19

34. Soares Cajaiba-Soares, A. M., Martinez-Silveira, M. S., Paim Miranda, D. L., de Cássia Pereira Fernandes, R., & Reis, M. G. (2021). Healthcare Workers’ Knowledge about Chagas Disease: A Systematic Review. The American Journal of Tropical Medicine and Hygiene, 104(5), 1631–1638. 10.4269/ajtmh.20-1199

35. Sosa-Estani, S., Cura, E., Velazquez, E., Yampotis, C., & Segura, E. L. (2009). Etiological treatment of young women infected with Trypanosoma cruzi, and prevention of congenital transmission. Revista da Sociedade Brasileira de Medicina Tropical, 42(5), 484–487. 10.1590/s0037-86822009000500002

36. Stimpert, K. K., & Montgomery, S. P. (2010). Physician awareness of Chagas disease, USA. Emerging Infectious Diseases, 16(5), 871–872. 10.3201/eid1605.091440

37. Tan, T. Q. (Ed.). (2023). American Trypanosomiasis: (Chagas Disease). In Red Book Atlas of Pediatric Infectious Diseases (p. 0). American Academy of Pediatrics. 10.1542/9781610026314-154

38. U.S. Census Bureau. (2021). Connecticut: 2020 Census, Connecticut’s Population Inched Up 0.9% Last Decade. Census.Gov. https://www.census.gov/library/stories/state-by-state/connecticut-population-change-between-census-decade.html

39. Verani, J. R., Montgomery, S. P., Schulkin, J., Anderson, B., & Jones, J. L. (2010). Survey of Obstetrician-Gynecologists in the United States About Chagas Disease. The American Journal of Tropical Medicine and Hygiene, *83*

40. Alarcón, A., Morgan, M., Montgomery, S. P., Scavo, L., Wong, E. C. C., Hahn, A., & Jantausch, B. (2016). Diagnosis and Treatment of Congenital Chagas Disease in a Premature Infant. Journal of the Pediatric Infectious Diseases Society, *5*(4), e28–e31. 10.1093/jpids/piw043

